# Hypertension, Insulin Resistance and Right Ventricular Dysfunction: Insights from a Single-Center Study and Mendelian Randomization Analysis

**DOI:** 10.1101/2025.07.09.25331231

**Authors:** Yu Pan, Tingting Fu, Minghui Gong, Qiaobing Sun, Xu Zhao, Shuang Yin, Tao Cong, Yinong Jiang, Yan Liu

**Author notes:** **Corresponding author:** Yan Liu, Department of Cardiology, First Affiliated Hospital of Dalian Medical University, Dalian, Liaoning, 116011, China., Yinong Jiang, Department of Cardiology, First Affiliated Hospital of Dalian Medical University, Dalian, Liaoning, 116011, China., Tao Cong, Department of Cardiology, First Affiliated Hospital of Dalian Medical University, Dalian, Liaoning, 116011, China. **First authorship:** Yu Pan and Tingting Fu have contributed equally to this work and share first authorship.

## Abstract

**Background:** Right ventricular (RV) dysfunction is increasingly recognized in hypertensive patients, often linked to metabolic abnormalities. The triglyceride glucose-body mass index (TyG-BMI), a marker of insulin resistance (IR), has been associated with cardiovascular outcomes, but its role in RV dysfunction remains unclear.

**Objectives:** This study aimed to evaluate the association between TyG-BMI and right ventricular free wall longitudinal strain (RVFWLS) in hypertensive patients using two-dimensional speckle tracking echocardiography. Key risk factors influencing RVFWLS were identified, and the contributions of TyG-BMI and its components-fasting plasma glucose (FPG), triglycerides (TG), and BMI-were examined. Causality was verified via multivariable Mendelian randomization (MVMR).

**Methods:** A total of 280 hypertensive patients underwent echocardiographic and clinical assessments. RVFWLS was used to evaluate RV function, with -21% as the threshold for normal. Predictor of RVFWLS were identified using LASSO regression and random forest models. Multiple linear regression and mediation analysis were performed to assess the direct and indirect effects of TyG-BMI on RVFWLS, while weighted quantile sum regression was conducted to quantify the contributions of TyG-BMI components. MVMR was utilized to explore causal relationships between TyG-BMI components, hypertension, and RV ejection fraction (RVEF).

**Results:** TyG-BMI emerged as the strongest predictor of RVFWLS, followed by HDL-C, SBP, hs-CRP, heart rate, and diabetes history. TyG-BMI was independently associated with RVFWLS (β = 0.03, P < 0.001). Mediation analysis revealed hs-CRP partially mediated this association (indirect effect: 0.002; 95% CI: 0.001–0.001; P=0.02). Among TyG-BMI components, FPG contributed most to RVFWLS (50%), followed by BMI (33%) and TG (17%). MVMR confirmed a causal role of FPG in reduced RVEF (P < 0.05).

**Conclusion:** TyG-BMI is strongly associated with subclinical RV dysfunction in hypertension, with hs-CRP acting as a partial mediator and FPG as the dominant contributor.

**Clinical Perspective:** *What is new?:*

- This study identifies TyG-BMI as a novel and robust predictor of right ventricular free-wall longitudinal strain (RVFWLS) in hypertensive patients, emphasizing its significance as a marker of metabolic and cardiovascular health.
- Among TyG-BMI components, fasting plasma glucose (FPG) emerges as the strongest contributor to RV dysfunction, supported by clinical data and Mendelian randomization analysis.

*What are the clinical implications?:*

- Assessing TyG-BMI can help clinicians identify hypertensive patients at higher risk for subclinical RV dysfunction.
- Early metabolic interventions targeting FPG and systemic inflammation may prevent or mitigate RV remodeling progression, minimizing further cardiovascular complications.

## Introduction

Hypertension, one of the most prevalent chronic conditions worldwide, imposes a significant public health burden due to its high morbidity and mortality. Hypertension-induced target organ damage (TOD) represents the preclinical stage of cardiovascular disease, and correlates with an increased risk of future cardiovascular events. Left ventricular (LV) remodeling is the most well-studied manifestation of cardiac TOD and a key determinant of adverse outcomes in hypertensive patients(1). However, the impact of hypertension on right ventricle (RV) has received comparatively less attention. Advanced imaging has revealed early-stage subclinical RV dysfunction in hypertensive patients without LV impairment or pulmonary hypertension(2,3).

Hypertension commonly coexists with a cluster of metabolic abnormalities, including obesity, dyslipidemia, glucose intolerance, and hyperuricemia. Moreover, RV remodeling tends to be more pronounced when hypertension is accompanied by these metabolic disorders(4). Metabolic abnormalities exacerbate RV remodeling. Recent studies have also demonstrated that RV remodeling exerts an independent impact on cardiovascular performance, exercise capacity, and adverse outcomes in hypertensive patients(3,5). This highlights the need to understand the mechanisms linking metabolic disorders with RV dysfunction in hypertensive patients.

Insulin resistance (IR) plays an important role in this interplay, serving as a key mediator of metabolic and cardiovascular dysfunction. The triglyceride-glucose (TyG) index is a widely used surrogate marker of IR(6). However, the TyG-body mass index (TyG-BMI), which builds upon the TyG index by incorporating BMI to account for adiposity-related metabolic burden, has emerged as a more robust indicator of IR(7). TyG-BMI has already demonstrated strong predictive values in the incidence of hypertension(8,9) and cardiovascular outcomes(10–14). However, no clinical studies to date have specifically investigated the relationship between TyG-BMI and RV function in hypertensive patients.

The effects of IR on cardiac remodeling are multifaceted, with low-grade systemic inflammation emerging as a critical mediator linking metabolic dysregulation to myocardial remodeling(15–17). Systemic inflammation, reflected by hs-CRP, correlates with CVD (18,19). However, their potential role as intermediaries between metabolic stressors like the TyG-BMI index and RV function remains unclear. Identifying and quantifying such mediation pathways is crucial for understanding more deeply the mechanisms underlying RV remodeling in hypertensive patients.

Additionally, while the TyG-BMI’s predictive utility in cardiovascular outcomes is well-established, the internal contributions of its individual components, fasting plasma glucose (FPG), triglycerides (TG), and BMI, to myocardial remodeling, especially for the RV, remain unexplored.

Therefore, the primary objective of this study is to assess the relationship between TyG-BMI and RV free-wall longitudinal strain (RVFWLS), a sensitive and early marker of RV dysfunction(20), in patients with primary hypertension. Specifically, this study aims to explore both direct effects and potential mediation pathways linking the TyG-BMI index to RVFWLS, with a focus on systemic inflammatory biomarkers such as hs-CRP and internal contributions of TyG-BMI components (TG, FPG, and BMI).

To achieve these objectives, an integrative approach combining clinical observational data, machine learning techniques for variable selection and weight assessment, as well as Mendelian randomization (MR) analysis for causal inference, was employed. This multi-level framework provides novel insights into the metabolic stressors contributing to RV remodeling in hypertension.

## Methods

The studies involving human participants were reviewed and approved by the Ethics Committee of the First Affiliated Hospital of Dalian Medical University (Approval Number: PJ-KS-KY-2023-530). The patients/participants provided their written informed consent to participate in this study. Written informed consent was obtained from the individual(s) for the publication of any potentially identifiable images or data included in this article.

### Study Design

This study combines a clinical observational analysis of RVFWLS in hypertensive patients with a multivariable Mendelian randomization (MVMR) examining causal links between metabolic traits and RV ejection fraction (RVEF). The overall study workflow is presented in Figure 1.

**Figure 1.**
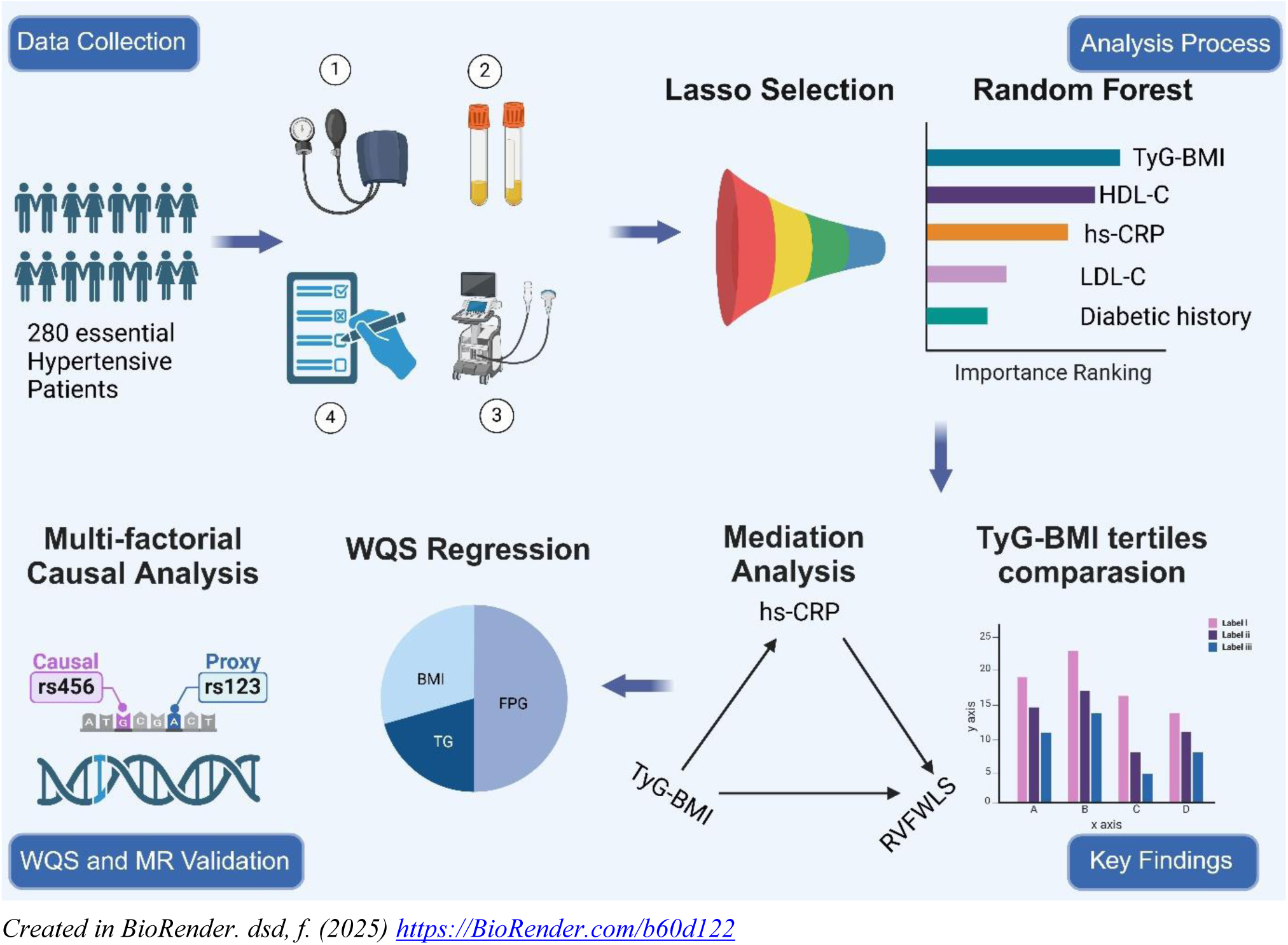
Graphical Abstract: Study Workflow and Key Findings

#### 1. Clinical Research

##### 1.1 Study Population

This study included 280 patients with primary hypertension who were hospitalized at the Hypertension and Heart Failure Department of the First Affiliated Hospital of Dalian Medical University between October 2021 and June 2023. Patients were eligible if they were aged 18–85 years and had a confirmed diagnosis of primary hypertension(20) following comprehensive assessments, including physical examination, blood biochemistry tests, secondary hypertension screening, and transthoracic echocardiography, and had clear cardiac strains images stored.

The exclusion criteria included:

(1) suspected or confirmed secondary hypertension; (2) pulmonary diseases or pulmonary artery hypertension caused by pulmonary embolism; (3) history of angina, myocardial infarction, coronary artery revascularization surgery, or coronary stenosis ≥50% on cardiac CT angiography; (4) heart failure with reduced LV ejection fraction (LVEF <50%); (5) moderate or severe valvular stenosis or regurgitation; (6) severe hepatic or renal dysfunction; (7) atrial fibrillation or flutter; (8) malignancies; and (9) poor echocardiographic image quality.

##### 1.2 Data Collection

Clinical data, including demographics (age, height, weight, and sex), blood pressure measurements, and histories of diabetes mellitus, hypertension, smoking, alcohol use, and antihypertensive medication use, were retrieved from electronic medical records. Blood pressure was measured using a calibrated sphygmomanometer.

Venous fasting blood samples were collected in the early morning after at least 12 hours of fasting to assess FPG, glycated hemoglobin A1c (HbA1c), serum creatinine (Scr), total cholesterol (TC), TG, high-density lipoprotein cholesterol (HDL-C), low-density lipoprotein cholesterol (LDL-C), hs-CRP, and uric acid (UA). The TyG-BMI was derived by multiplying TyG index (ln[TG×FPG/2]) by BMI.

##### 1.3 Echocardiographic Assessment

Echocardiography was performed using standard Vivid E9 equipment (GE Vingmed Ultrasound). Imaging followed the 2018 recommendations of the European Society of Cardiology(20).

LV parameters included end-diastolic diameter (LVEDD), posterior wall thickness at end-diastole (LVPWTD), interventricular septum thickness (IVSTD), and ejection fraction (LVEF), which was measured using Simpson’s biplane method. LV mass index (LVMI) was calculated using the Devereux formula. The left atrial volume index (LAVI) and mitral inflow velocity-to-late diastolic velocity ratio (E/A) were also measured to evaluate atrial size and diastolic function, respectively. In addition, the ratio of early diastolic filling velocity to mitral annular early diastolic velocity (E/e’) was calculated as a surrogate marker of left ventricular filling pressure, providing further insight into diastolic function. RV parameters included RV outflow tract diameter (RVOTD), end-diastolic diameter (RVEDD), tricuspid annular plane systolic excursion (TAPSE), RV fractional area change (RVFAC), tricuspid annular velocity during systole (S’), and RVEF.

Advanced myocardial strain analysis was performed using two-dimensional speckle-tracking echocardiography (2D-STE). Focused four-chamber apical views were used to calculate RVFWLS (Supplementary Figure 1). LV global longitudinal strain (LVGLS) was obtained by analyzing standard apical views of the LV including four-chamber, two-chamber, and three-chamber planes. All strain measurements were verified offline by independent researchers using EchoPAC software version 202 (GE Vingmed Ultrasound). Any erroneous myocardial tracking was either corrected manually or excluded from the analysis.

Intra- and inter-observer agreements for LVGLS and RVFWLS were evaluated using intraclass correlation coefficients (ICCs) with 95% confidence intervals (95% CI). The results, summarized in Supplement Table 2, showed excellent intra-observer reliability and good-to-excellent inter-observer reliability for both LVGLS and RVFWLS.

##### 1.4 Statistical Analysis

Continuous data were assessed by Shapiro-Wilk test and compared using ANOVA (or Welch’s ANOVA if unequal variances), followed by least significant difference (LSD) test for pairwise comparisons if the overall P value was <0.05. For non-normally distributed variables, the Kruskal-Wallis test was employed, followed by pairwise comparisons using the Bonferroni correction method when P < 0.05. Categorical variables were reported as frequencies and percentages and analyzed using chi-squared tests or Fisher’s exact test, as appropriate.

To identify risk factors influencing RVFWLS, LASSO regression was employed to reduce multicollinearity among predictors by penalizing irrelevant coefficients toward zero. The target variable for this analysis was RVFWLS, dichotomized as ≤ -21% for abnormal function based on guidelines from the Chinese Society of Echocardiography(21). The variables included in the model for selection were: sex, age, BMI, duration of hypertension, history of diabetes, history of obstructive sleep apnea-hypopnea syndrome (OSAS), smoking history, alcohol consumption history, medication history, LVEF, LVGLS, LVMI, HR, SBP, DBP, heart rate, BNP, HbA1c, FPG, UA, Scr, TC, TG, HDL-C, LDL-C, TyG, TyG-BMI, eGFR, and hs-CRP. Variables retained after LASSO regression were ranked using a random forest analysis to measure their importance in predicting RVFWLS.

Given that the TyG-BMI index emerged as the top-ranking predictor in the random forest analysis, patients were divided into tertiles based on TyG-BMI levels for comparisons. Univariate and multivariate linear regression analyses were performed to evaluate associations between TyG-BMI and echocardiographic parameters. Mediation analysis using the mediation package in R was conducted to examine whether hs-CRP or LVGLS mediated the relationship between TyG-BMI and RVFWLS. Weighted quantile sum (WQS) regression further analyzed the contribution of individual components of TyG-BMI (FPG, TG, and BMI) to RVFWLS.

#### 2. Multivariable Mendelian Randomization

##### 2.1 Data Sources

Mendelian randomization analyses were conducted to assess causal relationships between clinical metabolic traits and RVEF. Exposure data for TG (ieu-a-302), FPG (ieu-b-114), BMI (ukb-b-19953), and hypertension (ukb-b-12493) were retrieved from publicly available genome-wide association studies in the IEU OpenGWAS project. RVEF outcome data (GCST90134591) were obtained from GWAS Catalog.

##### 2.2 Instrumental Variable Selection

Single nucleotide polymorphisms (SNPs) significantly associated with exposure traits (P<5×10^-8^) were selected as instrumental variables. Linkage disequilibrium pruning was performed using thresholds (clump_kb=10000; clump_r^2^=0.001) to remove redundant variants within linkage blocks. Harmonization ensured alignment of alleles across exposures and outcomes.

##### 2.3 Causal Effect Estimation

Causal relationships were estimated using inverse variance weighting (IVW) as the primary method. Horizontal pleiotropy effects were evaluated via MR-Egger regression, while weighted median estimation provided robust causal effect estimates even when some instruments failed validity assumptions. Sensitivity analyses were conducted to assess the robustness of the results. Heterogeneity among instrumental variables was evaluated using Cochran’s Q test, while instrument strength was assessed via conditional F-statistics. Potential violations from outlier variants or heterogeneity were further examined using the MR-PRESSO global test, which identifies and corrects for outliers when applicable.

All statistical analyses were performed using SPSS (version 26), R software (version 4.4.2) and RStudio (version 2024.9.1.394). The LASSO regression was implemented using the “glmnet” package, random forest analysis using “randomForest”, mediation analysis using “mediation”, and WQS regression using “gWQS”. For the Mendelian randomization analyses, we used the “TwoSampleMR” package for primary analyses and the “MVMR” package for multivariable MR analyses. The “TwoSampleMR” R package with integrated PLINK functionality was employed for LD pruning. The “MR-PRESSO” package was used for pleiotropy assessment. Statistical significance was set at P < 0.05 for all analyses.

## Results

### LASSO Regression for Screening Predictive Factors

In this study, predictive factors were identified using LASSO regression (Figure 2). The path plot (Figure 2A) showed that as the penalty parameter λ decreased, variables were gradually included in the model. When λ = 0.031606, the cross-validation curve (Figure 2B) exhibited the minimum mean error, resulting in the selection of six significant predictors: TyG-BMI index, hs-CRP, HDL-C, SBP, heart rate (HR), and a history of diabetes.

**Figure 2.**
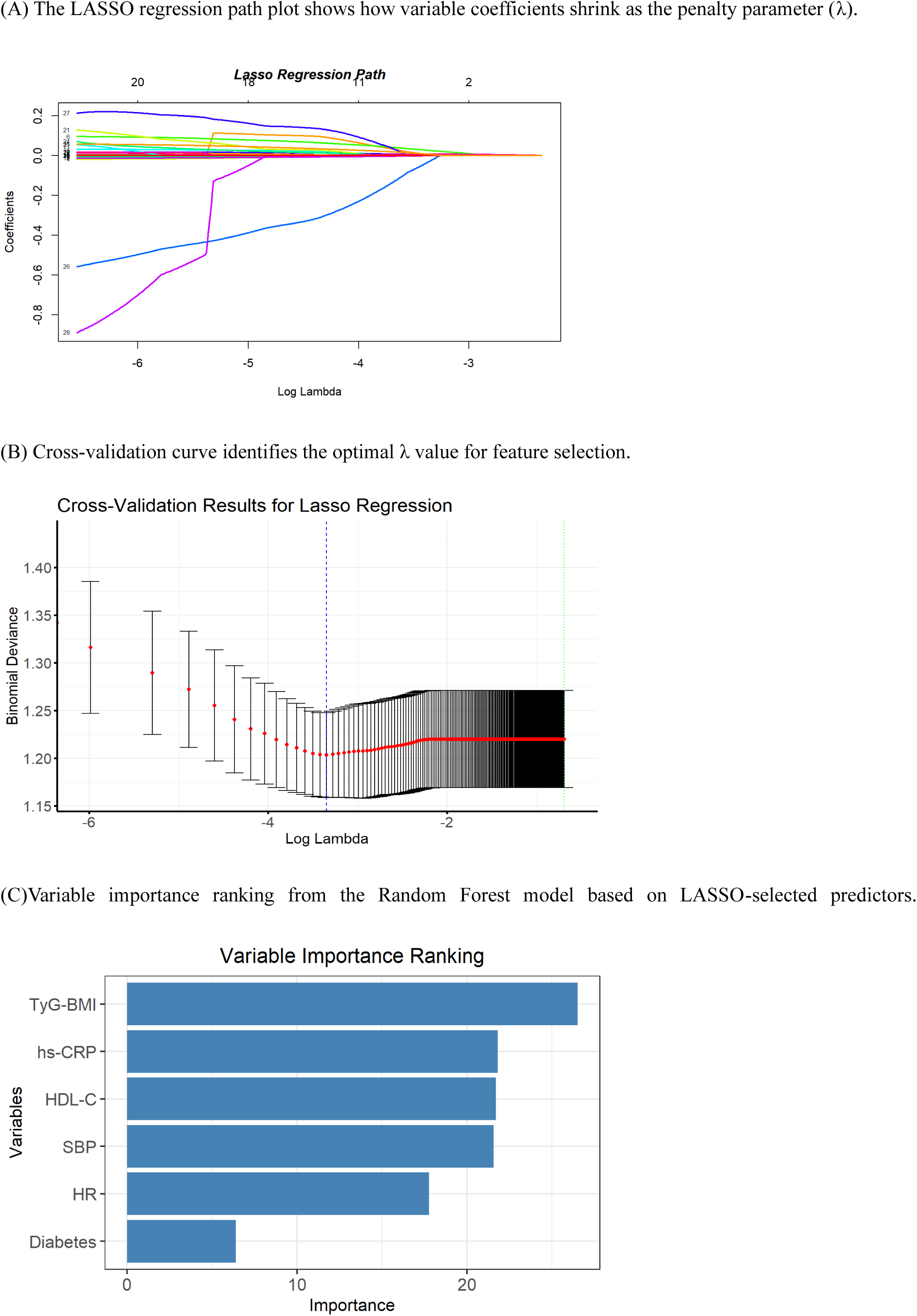
Selection and Ranking of Predictive Factors for RVFWLS

### Ranking Variable Importance Using Random Forest Model

The random forest model ranked the importance of the six factors identified by LASSO regression. The importance scores were as follows: TyG-BMI index (26.51), HDL-C (21.82), SBP (21.70), hs-CRP (21.57), HR (17.76), and history of diabetes (6.39) (Figure 2C).

### Comparison of Clinical and Biochemical Characteristics Across TyG-BMI Tertiles

Participants were divided into three groups based on TyG-BMI index tertiles (Tertile 1, Tertile 2, Tertile 3) with mean indices of 191 ± 21.0, 234 ± 10.0, and 278 ± 20.3, respectively. Table 1 summarizes the demographic and clinical characteristics across tertiles. Significant differences were noted among groups (P < 0.05) in age, sex, smoking status, history of OSAS, BMI, heart rate, FPG, UA, Scr, eGFR, BNP, TG, TyG index, HDL-C, LDL-C, and hs-CRP. Participants in Tertile 3 exhibited worse metabolic profiles compared to Tertile 1, including higher BMI, FPG, and LDL-C values but lower HDL-C (P < 0.05).

**Table 1.** Comparison of Baseline Characteristics among Tertiles of the TyG-BMI Index.

|  | **Tertile1**<br>**N=93** | **Tertile2**<br>**N=94** | **Tertile3**<br>**N=93** | **p** |
| --- | --- | --- | --- | --- |
| Age(years) | 54.8±15.4 | 56.3±13.0 | 50.5±13.8<sup>†</sup> | 0.015 |
| Male (%) | 38 (40.9%) | 47 (50.0%) | 59 (63.4%)* | 0.008 |
| BMI (kg/m2) | 23.1±2.31 | 26.7±1.58** | 30.1±2.37**† | <0.001 |
| Smoke (%) | 19 (20.4%) | 27 (28.7%) | 37 (39.8%)<sup>†</sup> | 0.015 |
| Drink (%) | 11 (11.8%) | 13 (13.8%) | 22 (23.7%) | 0.066 |
| Diabetes (%) | 13 (14.0%) | 21 (22.3%) | 22 (23.7%) | 0.201 |
| OSAS (%) | 6 (6.38%) | 13 (14.0%) | 21 (22.6%)* | 0.007 |
| Hypertension-Duration(years) | 4.00 (1.00, 13.0) | 5.00 (2.00, 14.8) | 6.00 (2.00, 13.0) | 0.384 |
| SBP (mmHg) | 157±23.0 | 160±21.1 | 160±22.9 | 0.524 |
| DBP (mmHg) | 92.9±17.6 | 95.4±18.2 | 96.7±16.0 | 0.319 |
| HR (bpm) | 69.5 (63.0,74.0) | 70.0 (64.0,77.0) | 73.0 (67.0,80.0)* | 0.013 |
| FPG (mmol/l) | 4.67 (4.32, 5.09) | 5.08 (4.64, 5.78)** | 5.26 (4.77, 6.58)** | <0.001 |
| HbA1c (%) | 5.99±1.54 | 6.25±1.09 | 6.26±1.39 | 0.295 |
| UA (umol/l) | 333 (276, 380) | 370 (290, 432)* | 401 (344, 465)**† | <0.001 |
| Scr (μmol/l) | 60.0 (54.0, 71.0) | 63.0 (52.2, 84.8) | 68.0 (58.0, 85.0)**† | 0.025 |
| eGFR(90 mL/min/1.73m<sup>2</sup>) | 80.6 (66.6, 94.2) | 85.7 (67.0,104) | 95.2 (76.8,114)**† | 0.003 |
| BNP(pg/mL ) | 27.4 (15.5, 57.3) | 23.4 (14.7, 55.5) | 18.1 (8.98, 52.7)* | 0.023 |
| TC (mmol/l) | 4.65 (4.05, 5.35) | 4.72 (4.22, 5.38) | 4.96 (4.09, 5.79) | 0.149 |
| TG (mmol/l) | 0.98 (0.80, 1.24) | 1.46 (1.18, 1.82)** | 2.07 (1.51, 3.17)**† | <0.001 |
| TyG | 8.27±0.44 | 8.77±0.47** | 9.26±0.76**† | <0.001 |
| TyG-BMI | 191±21.0 | 234±10.0** | 278±20.3**† | <0.001 |
| HDL-C (mmol/l) | 1.25 (1.11, 1.47) | 1.08 (0.92, 1.22)** | 1.07 (0.88, 1.22)** | <0.001 |
| LDL-C (mmol/l) | 2.52±0.69 | 2.69±0.67 | 2.79±0.75* | 0.030 |
| hs-CRP (mg/L) | 0.69 (0.31, 1.61) | 1.04 (0.45, 1.76) | 1.49 (0.81, 3.74)**† | <0.001 |
| Antihypertensive drugs |  |  |  |  |
| CCB | 42 (45.2%) | 55 (58.5%) | 60 (65.2%)* | 0.020 |
| ACEI/ARB/ARNI | 37 (39.8%) | 38 (40.4%) | 51 (54.8%) | 0.065 |
| β-blocker | 17 (18.3%) | 25 (26.6%) | 25 (26.9%) | 0.295 |
| Diuretic | 9 (9.68%) | 12 (12.8%) | 10 (10.8%) | 0.792 |

Note: BMI: Body Mass Index; OSAS:Obstructive Sleep Apnea Syndrome; SBP: Systolic Blood Pressure; DBP: Diastolic Blood Pressure; HR: Heart Rate; FPG: Fasting Plasma Glucose; HbA1c: Glycated Hemoglobin; UA: Serum Uric Acid; Scr: Serum Creatinine; eGFR: Estimated Glomerular Filtration Rate; BNP: B-type Natriuretic Peptide; TC: Total Cholesterol; TG: Triglycerides; TyG index: Triglyceride-Glucose Index; TyG-BMI index: Triglyceride-Glucose Body Mass Index; HDL-C: High-Density Lipoprotein Cholesterol; LDL-C: Low-Density Lipoprotein Cholesterol; hs-CRP: high-sensitivity C-Reactive Protein; CCB: Calcium Channel Blockers; ACEI: Angiotensin-Converting Enzyme Inhibitors; ARB: Angiotensin II Receptor Blockers; ARNI: Angiotensin Receptor-Nepriylsin Inhibitors; β-blocker: Beta Blockers; diuretic: Diuretics
\*p < 0.05, \*\*p < 0.001 vs. the Tertile 1; †p < 0.05, ††p < 0.001 vs. the Tertile 2

### Comparison of Echocardiography Parameters Across TyG-BMI Tertiles

Echocardiography parameters comparing groups by TyG-BMI tertiles are presented in Table 2. For LV parameters, LVEDD and LVMI were significantly higher in Tertile 3 compared to Tertile 1 (P < 0.05). LVGLS was significantly lower in Tertile 2 and Tertile 3 compared to Tertile 1 (P < 0.05), with a pronounced reduction in Tertile 3 compared to Tertile 2 (P < 0.05). Regarding diastolic function, E/A was significantly decreased in Tertile 2 compared to Tertile 1 (P < 0.05); however, no significant differences were observed between Tertile 3 and the other two groups. E/e’ were comparable among three groups (P > 0.05).

**Table 2.** Impact of the TyG-BMI Index on Biventricular Structure and Function.

|  | **Tertile1** | **Tertile2** | **Tertile3** | **P** |
| --- | --- | --- | --- | --- |
|  | **N=93** | **N=94** | **N=93** |  |
| LVEDD | 46.0 (44.0, 49.0) | 47.0 (45.0, 49.0)* | 48.0 (46.0, 51.0)** | <0.001 |
| LVMI | 92.8 (77.0, 109) | 101 (87.6, 112) | 105 (86.6, 117)* | 0.014 |
| E/A | 0.90 (0.75, 1.15) | 0.78 (0.69, 0.93)* | 0.81 (0.69, 1.06) | 0.042 |
| E/e' | 9.00 (7.00, 10.6) | 9.00 (7.78, 11.0) | 9.00 (7.00, 11.3) | 0.648 |
| EDT | 192 (167, 224) | 212 (190, 244)* | 203 (173, 230) | 0.003 |
| LVEF | 59.0 (55.0, 64.0) | 59.0 (56.0, 62.8) | 59.0 (55.0, 63.0) | 0.590 |
| LVGLS | -17.64±3.17 | -17.56±3.03** | -15.85±3.20**† | <0.001 |
| RVEDD | 17.0 (16.0, 18.0) | 17.0 (16.0, 18.0) | 18.0 (17.0, 19.0)**† | <0.001 |
| TAPSE | 19.0 (18.0, 22.0) | 19.0 (17.0, 22.0) | 20.0 (17.0, 22.0) | 0.621 |
| S' | 13.0 (11.0, 15.0) | 12.0 (11.0, 15.0) | 13.0 (12.0, 14.0) | 0.603 |
| RVFAC | 0.50±0.11 | 0.49±0.12 | 0.49±0.11 | 0.573 |
| RVEF | 70.0 (64.0, 76.0) | 70.0 (60.2, 76.0) | 67.0 (57.0, 75.0) | 0.247 |
| RVFWLS | -20.36±4.05 | -19.26±4.47 | -17.50±3.77**†† | <0.001 |

Note: LVEDD: Left Ventricular End-Diastolic Diameter; LVMI: Left Ventricular Mass Index; E/A: Ratio of Early (E) to Late (A) Ventricular Filling Velocities; E/e': Ratio of Early Diastolic Mitral Inflow Velocity to Early Diastolic Mitral Annular Velocity; EDT: Deceleration Time of Early Diastolic Mitral Inflow Velocity; LVEF: Left Ventricular Ejection Fraction; GLS: Global Longitudinal Strain; RVEDD: Right Ventricular End-Diastolic Diameter; TAPSE: Tricuspid Annular Plane Systolic Excursion; S': Tricuspid Annular Systolic Velocity; RVFAC: Right Ventricular Fractional Area Change; RVEF: Right Ventricular Ejection Fraction; RVFWLS: Right Ventricular Free Wall Global Longitudinal Strain.
\*p < 0.05, \*\*p < 0.001 vs. Tertile1; †p < 0.05, ††p < 0.001 vs. Tertile2

For RV parameters, RVEDD was significantly higher in Tertile 3 compared to Tertile 1 (P < 0.05). RVFWLS was lower in both Tertile 2 and Tertile 3 compared to Tertile 1 (P < 0.05), with more marked reduction in Tertile 3 than in Tertile 2 (P < 0.05). The results of the group comparison for RVFWLS and RVEDD are further visualized in Figure 3.

**Figure 3.**
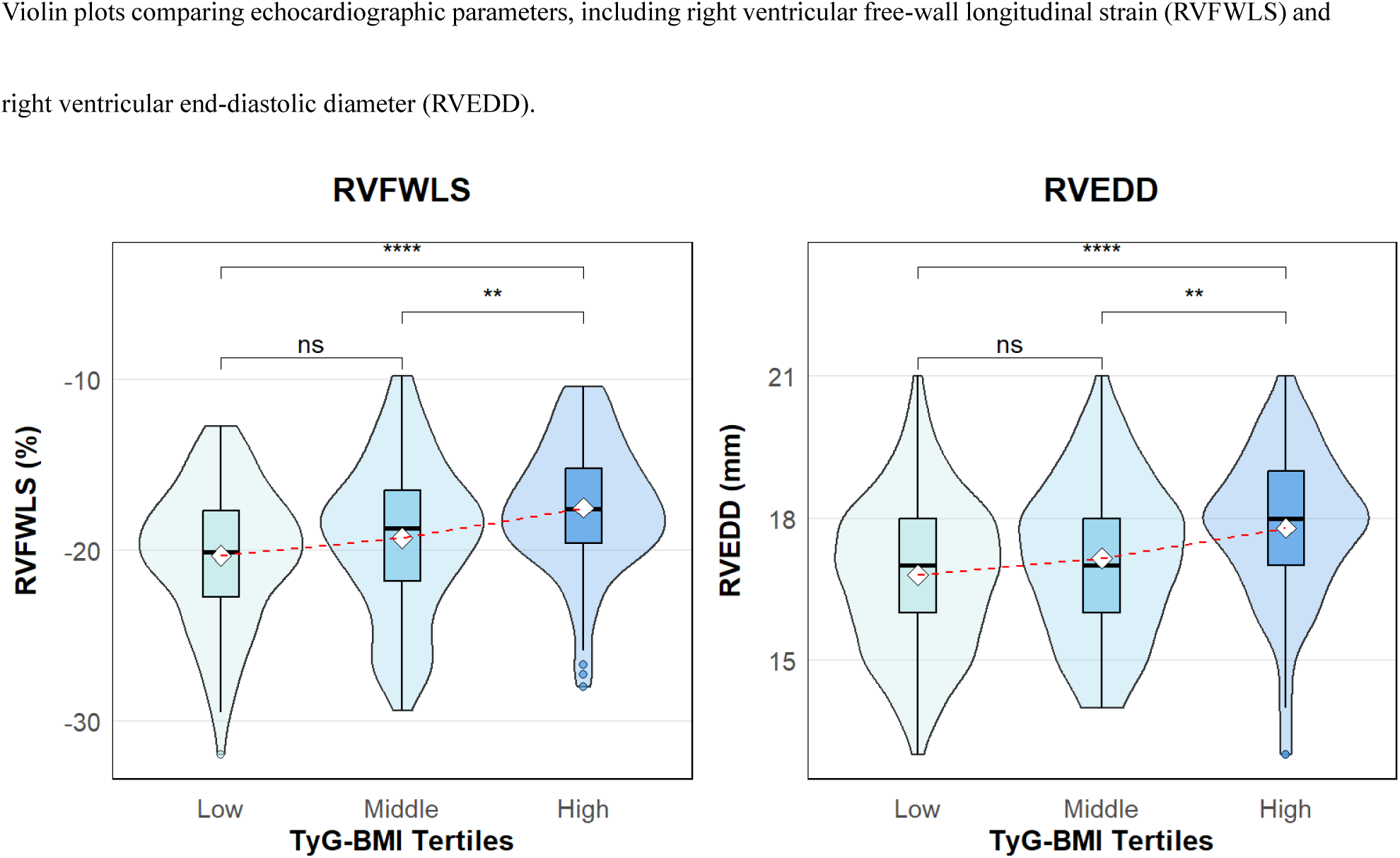
Group Comparisons of Echocardiographic Parameters Across TyG-BMI Tertiles

### Linear Regression Analysis of TyG-BMI Index on Bi-Ventricular Structure and Function

Linear regression analysis demonstrated significant associations between the TyG-BMI index and RV parameters, including RVEDD (0.01 [95% CI: 0.01–0.02], P < 0.001) and RVFWLS (0.03 [95% CI: 0.02–0.04], P < 0.001) (Figure 4A). Similarly, significant associations were observed for LV parameters, including LVEDD (0.03 [95% CI: 0.02–0.05], P < 0.001), LVMI (0.12 [95% CI: 0.05–0.19], P < 0.001), and LVGLS (0.02 [95% CI: 0.01–0.03], P < 0.001).

**Figure 4.**
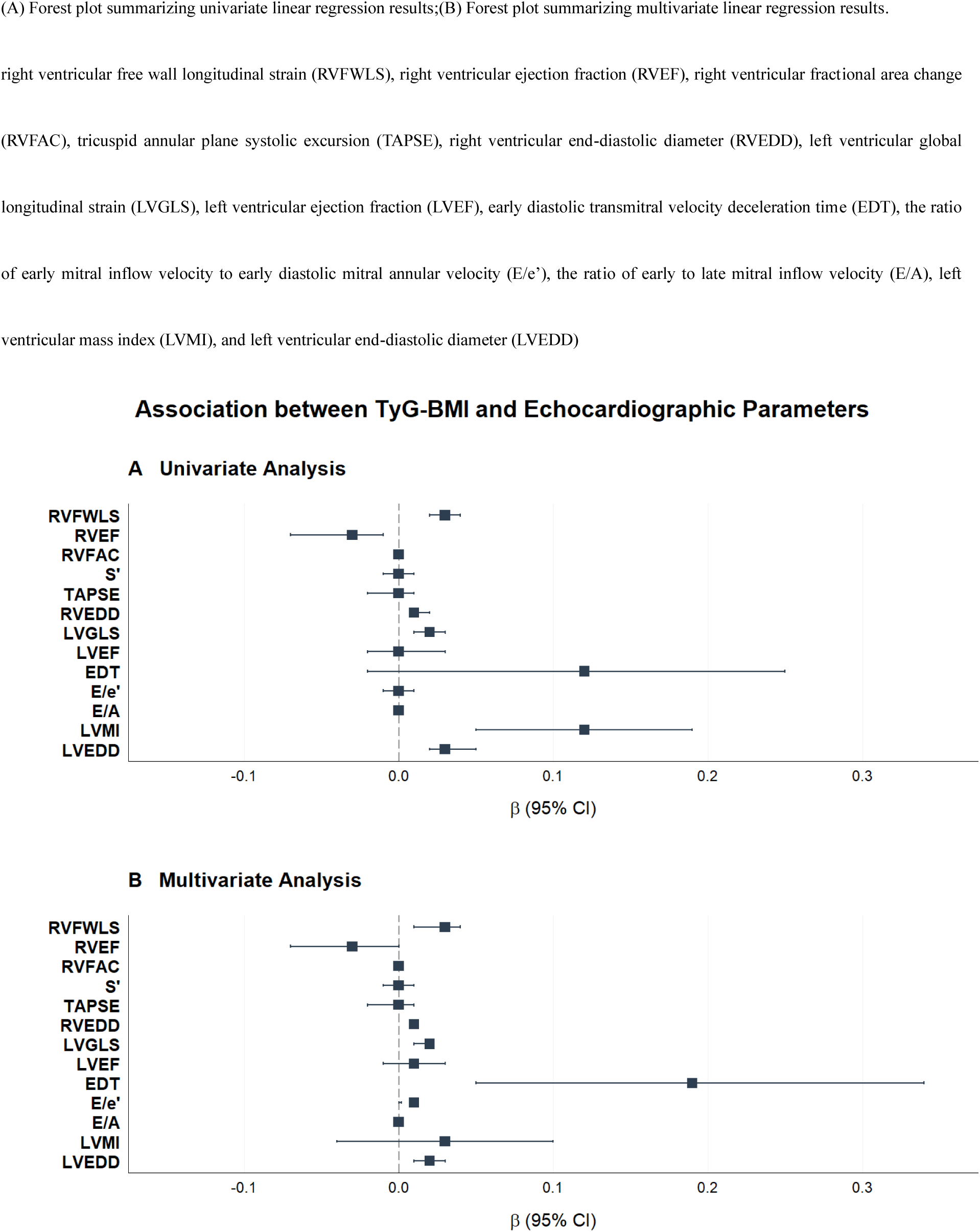
Univariate and Multivariate Linear Regression Results of TyG-BMI’s Association with Biventricular Echocardiographic Function

Multivariate regression analysis, adjusted for age, sex, diabetes, hypertension history, smoking history, alcohol consumption, SBP, DBP, heart rate (HR) and medication use (e.g., ACEI/ARB, β-blockers, diuretics, and CCB), further confirmed that the TyG-BMI index remained independently associated with RV parameters, including RVEDD (0.01 [95% CI: 0.01–0.01], P < 0.001) and RVFWLS (0.03 [95% CI: 0.01–0.04], P < 0.001), as shown in Figure 4B. Additionally, significantly independent associations were noted with LV parameters, including LVEDD (0.02 [95% CI: 0.01–0.03], P < 0.001), LVGLS (0.02 [95% CI: 0.01–0.02], P = 0.002), and EDT (0.19 [95% CI: 0.05–0.34], P = 0.008). Nonlinear evaluations using restricted cubic splines showed no evidence of a nonlinear relationship between TyG-BMI index and RV echocardiographic indices (RVFWLS and RVEDD) (P > 0.05) (Supplement Figure 2).

### Mediation Effect of hs-CRP on TyG-BMI Index and RVFWLS

Mediation analysis demonstrated an independent association between TyG-BMI index and RVFWLS through hs-CRP but not LVGLS (Supplement Table 1). The indirect effect of hs-CRP was statistically significant (effect size = 0.002; bootstrap CI: [0.001–0.010], P =0.02) (Figure 5), while LVGLS did not display mediation effects (effect size = 0.000; bootstrap CI: [0.000–0.000], P =1.00).

**Figure 5.**
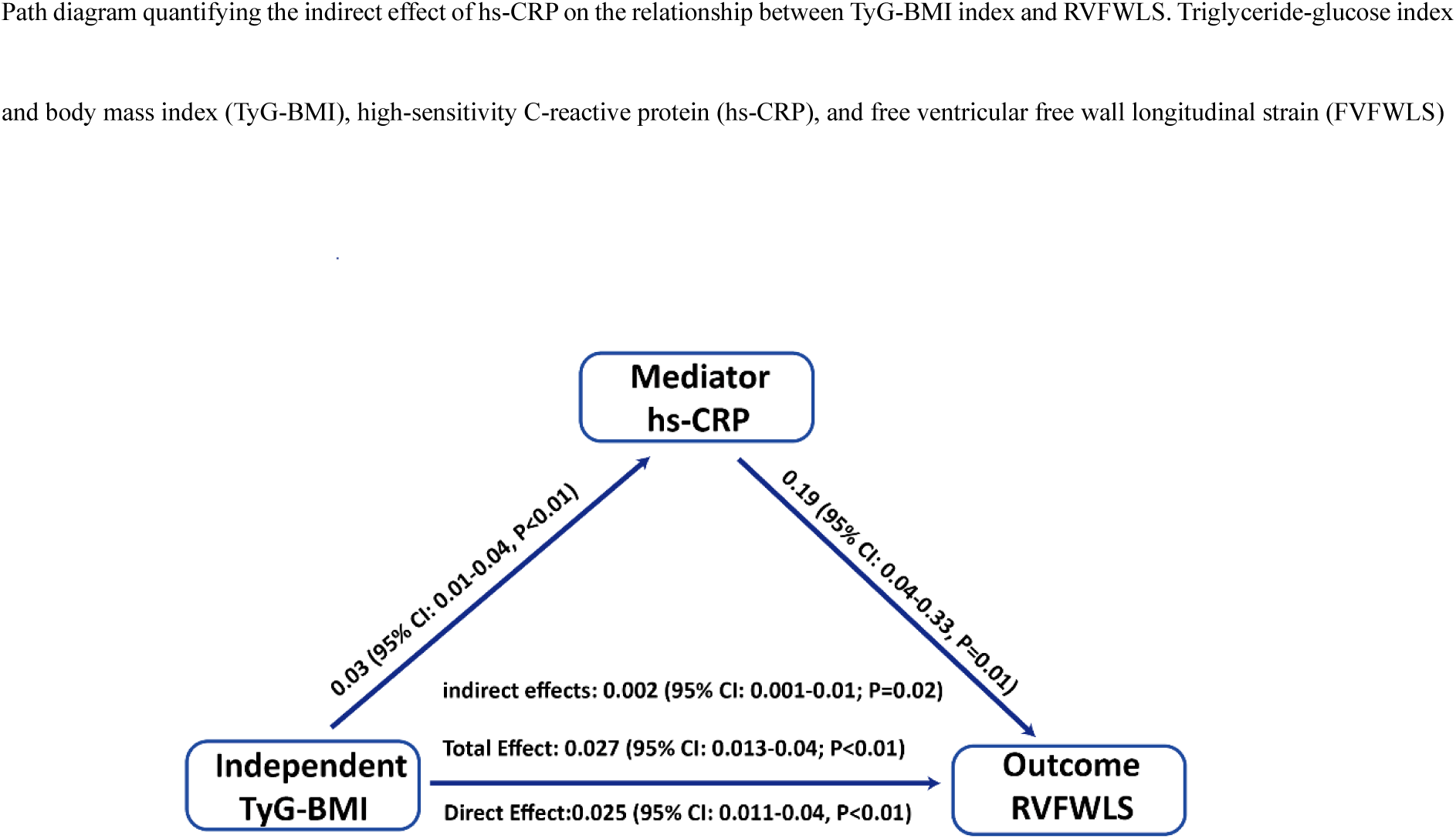
Mediation Effect of High-Sensitivity C-Reactive Protein (hs-CRP) in the Association Between TyG-BMI Index and RVFWLS

### WQS Validation of Component Weights for TyG-BMI Index

A WQS regression analysis was performed to confirm the contributions of TyG-BMI components (TG, FPG, BMI) to RVFWLS variance after adjusting for potential confounders, including age, sex, diabetes, hypertension history, smoking history, alcohol consumption, SBP, DBP, HR and medication use (e.g., ACEI/ARB, β-blockers, diuretics, and CCB) (Figure 6A). Among the components, FPG exhibited the highest weight (50%), followed by BMI (33%) and TG (17%).

**Figure 6.**
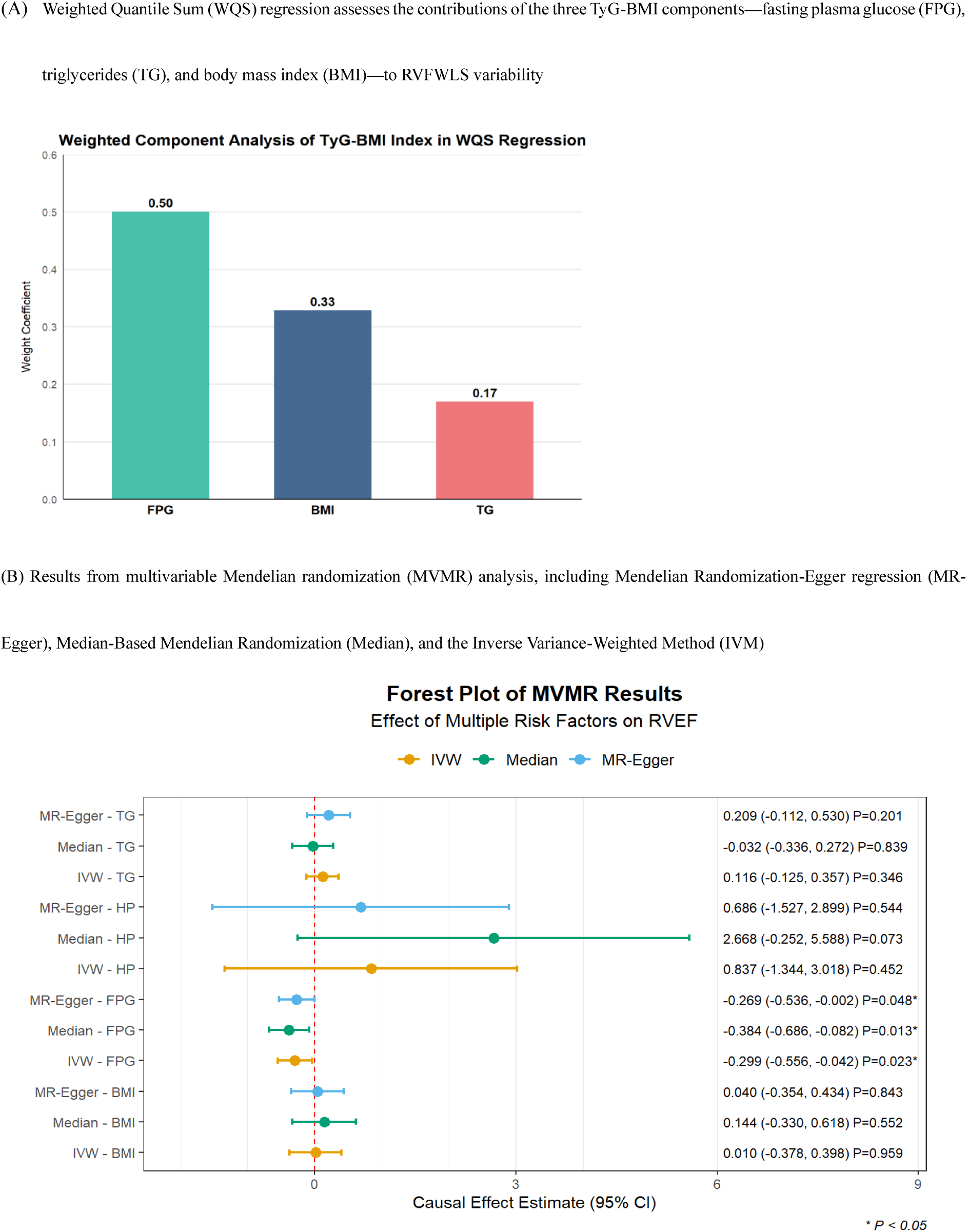
Combined Analysis of TyG-BMI Components and Their Causal Effects on Right Ventricular Function

### Multivariable MR Analysis of TyG-BMI Components on RVEF

Multivariable MR analysis further investigated the causal effects of TyG-BMI components (TG, FPG, BMI) and hypertension on RVEF using IVW analysis (Figure 6B). Only FPG exhibited a significant negative causal relationship with RVEF (β = -0.299; [95% CI: -0.557 to -0.042]; P = 0.023); TG (P = 0.346), BMI (P = 0.959), and hypertension (P = 0.452) showed no significant associations. MR-Egger regression supported the negative causal effect of FPG on RVEF while revealing no substantial horizontal pleiotropy bias (intercept P = 0.386). Weighted median estimation yielded consistent results, reinforcing the robustness of FPG’s negative causal association with RVEF (β = -0.384; [95% CI: -0.686 to -0.081]; P = 0.013).

Other supportive statistical analyses, including instrumental variable strength evaluation, heterogeneity tests, and MR-PRESSO global analysis, are detailed in the supplementary materials. In summary, FPG showed strong instrument validity (F-statistic >25), while hypertension exhibited relatively weaker instrument strength (F-statistic = 4.45). Heterogeneity assessments suggested variable genetic instruments (Q = 67.99; df = 40; P = 0.004), though no outlier instruments were identified despite MR-PRESSO detecting global heterogeneity (RSSobs = 85.17; P = 0.008).

## Discussion

This study aimed to evaluate the relationship between the TyG-BMI and RV dysfunction in hypertensive patients, specifically utilizing RVFWLS as the primary indicator of subclinical RV myocardial remodeling. Our findings demonstrated that TyG-BMI was a potent predictor of RVFWLS and that FPG played a dominant role among TyG-BMI components in driving RV dysfunction, with hs-CRP serving as a partial mediator. Moreover, MVMR further confirmed a genetic causal relationship between FPG and RV systolic function.

As we know, hypertension contributes to RV dysfunction predominantly through hemodynamic mechanisms. Persistent overload in pulmonary circulation contributes to RV structural and functional remodeling, typically following the LV remodeling. However, accumulating evidences have indicated that hypertension can affect RV structure and function in the early stages, even in the absence of overt LV involvement(2,22–24). Our current study corroborates these findings by demonstrating that hypertensive patients with preserved LV function still exhibit significant impairments in RVFWLS, emphasizing mechanisms beyond hemodynamic overload in the impact of hypertension on RV function. Chronic hypertensive stress can induce systemic metabolic and inflammatory alterations that further impair RV function(25).

IR plays a pivotal role in the occurrence, progression and prognosis of CVD(26–29). IR is commonly associated with hypertension, particularly when accompanied by other metabolic disorders such as obesity, dyslipidemia, and glucose intolerance(28). The TyG-BMI has been recognized as a well-established and widely utilized surrogate marker for IR(7). In the current study, patients in the highest TyG-BMI group exhibited the most pronounced metabolic disturbances. These individuals had significantly elevated FPG, TG, LDL-C, and UA, alongside reduced HDL-C. This profile underscores the heightened cardiometabolic risk associated with elevated TyG-BMI and its utility in identifying individuals with metabolic derangements. Studies have shown that TyG-BMI outperforms other indices, such as TyG alone or METS-IR, in predicting the risk and prognosis of CVD, making it a valuable tool in clinical settings(30,31). IR has also been reported to contribute to LV structural and functional remodeling(32), linking metabolic dysfunction with subclinical cardiac remodeling. Furthermore, Zhai et al.’s study(31) has revealed that TyG-BMI has the strongest correlation with LV hypertrophy (LVH) in hypertensive patients than the other IR surrogates. These findings emphasize the utility of TyG-BMI in evaluating cardiovascular risk beyond conventional measures. In this study, patients in the highest TyG-BMI group exhibited significantly increased LVEDD and LVMI, alongside reduced LVGLS, RVEDD and RVFWLS. Furthermore, TyG-BMI was identified as a significant determinant of RVFWLS, with partial mediation by inflammation (hs-CRP). This novel finding highlighted the importance of TyG-BMI in evaluating biventricular dysfunction, expanding its clinical relevance from LV remodeling to biventricular myocardial status.

Inflammation plays a pivotal role in the pathophysiology of CVD, acting as both a trigger and amplifier of structural and functional changes in the heart(33,34). In this study, IR-driven inflammation (reflected by elevated hs-CRP levels) contributed to RV remodeling in hypertensive patients. In hypertensive patients, inflammation contributes to cardiac remodeling through mechanisms such as endothelial dysfunction, oxidative stress, and cytokine-mediated myocardial injury(25,35,36). Elevated levels of inflammatory markers, such as hs-CRP, interleukins, and tumor necrosis factor-alpha (TNF-α), are strongly associated with adverse cardiovascular outcomes, including ventricular dysfunction and fibrosis(37,38). IR increases cardiovascular risk via inflammation and oxidative stress(35,39,40).

In cardiovascular metabolic diseases, this IR-inflammation nexus drives maladaptive remodeling, contributing to LV hypertrophy and systolic dysfunction. Emerging evidence suggests that the RV may be even more vulnerable to these mechanisms. The RV’s unique structure and hemodynamic characteristics predispose it to earlier and more pronounced damage from metabolic and inflammatory insults compared to the LV(41,42). Hypertension is well-known to induce LV structural and functional remodeling, elevating pulmonary circulation pressure and eventually causing RV dysfunction through hemodynamic mechanisms. However, in this study, LVGLS did not mediate the relationship between IR and RV systolic dysfunction. This finding may be attributed to the patient population being in a stage of cardiac function compensation with relatively preserved LV function, indicating that RV dysfunction may arise independently of overt LV impairment in the early stage of hypertension. Notably, hs-CRP partially mediated this relationship, underscoring the role of inflammation in linking metabolic dysfunction to RV remodeling. These results highlight the need to consider RV vulnerability in hypertensive patients with metabolic disorders, as early detection and intervention may mitigate progression to overt RV failure.

To further elucidate the specific contributions of TyG-BMI components to RV dysfunction, we employed WQS regression, which revealed that FPG had the highest weight (50%), followed by BMI (33%) and TG (17%). This highlights glucose regulation as the most critical driver of RV remodeling among the metabolic components in hypertensive patients. The application of WQS provides an advantage by addressing multicollinearity within TyG-BMI and enabling a detailed decomposition of its effects. Huo et al.’study(43) has found TG as the strongest predictor of stroke risk in middle-aged and older adults, followed by BMI and FPG. Jiang et al.’ study(44) has elucidated the relationship between TyG-BMI and coronary artery calcification in patients on maintenance hemodialysis, with BMI identified as the primary contributor.

The MVMR analysis provided robust evidence for a genetic causal relationship between FPG and impaired RVEF, while no significant causal effects were observed for TG, BMI, or essential hypertension. These findings highlight glucose dysregulation as an independent driver of RV systolic dysfunction, distinct from the hemodynamic burden typically associated with hypertension. FPG as an independent causal driver of RV dysfunction, underscoring the independent role of glucose dysregulation in driving RV systolic dysfunction at a genetic level. There is no causal relationship was observed between TG or BMI and RV function, likely reflecting their more complex or indirect contributions. These results underscore the clinical potential of targeted glycemic control interventions for improving RV function. Previous MR analysis has demonstrated a causal relationship between hypertension and LV structural and functional dysfunction(45), highlighting the role of hemodynamic overload in driving LV remodeling. This divergence underscores fundamental differences in the pathophysiological responses of the LV and RV to hypertension and its associated metabolic disturbances. Unlike the LV, which is directly exposed to systemic arterial pressure, the RV is affected indirectly. This may explain the absence of a direct causal relationship between hypertension and RV function observed in our MR analysis. RV may be more vulnerable to metabolic disturbances in the early stages of disease progression. These results underscore the importance of targeted metabolic interventions, particularly optimizing glucose control, to prevent or mitigate RV dysfunction in hypertensive patients with concurrent metabolic derangements.

### Clinical Implication

TyG-BMI integrates the synergistic effects of FPG, TG, and BMI, thereby providing a more comprehensive reflection of metabolic disorder burden. This composite index is particularly useful for clinical screening of high-risk patients. The MR Analysis isolates the causal effect of FPG on RV dysfunction, suggesting that hyperglycemia itself may impair right ventricular function through direct mechanisms and providing genetic evidence that targeted interventions (such as hypoglycemic therapy) may directly improve right ventricular function.

The RV’s heightened susceptibility to inflammation and IR underscores the importance of early and comprehensive cardiovascular risk assessment in hypertensive patients, especially combined with the other metabolic disorders. Healthy dietary patterns and novel hypoglycemic medications, such as Glucagon-like peptide-1 (GLP-1) receptor agonists and Sodium-glucose cotransporter 2 (SLTG2) inhibitors, have the potential in improving IR and inflammation(46–50) in addition to optimizing metabolic regulation. Future research should validate targeted anti-inflammatory and metabolic therapies to effectively prevent or reverse RV remodeling in this high-risk population.

### Strengths and Limitations

This study has several strengths, including the comprehensive evaluation of RV dysfunction using advanced echocardiographic techniques, integration of machine learning models for risk factor identification, and the application of MVMR to establish causality. The use of TyG-BMI as an index of IR adds practical value, as it is easily accessible and clinically relevant.

However, the study has limitations. First, its single-center, cross-sectional design limits the generalizability of our findings and precludes assessment of temporal relationships exposure and outcomes. Second, although hs-CRP serves as a general marker of systemic inflammation, it may not fully capture the complex inflammatory processes involved in myocardial remodeling. Future large-scale clinical studies incorporating additional inflammatory biomarkers, such as IL-6 and TNF-α, could provide deeper insights into the role of inflammation in RV dysfunction and uncover stronger effects. Additionally, while MR analysis provides valuable insights into causality, its reliability is dependent on the availability and quality of genetic instruments. Moreover, due to the unavailability of relevant SNPs for RVFWLS in current GWAS databases, we opted to assess RVEF as a feasible alternative.

## Conclusion

Hypertension is associated with RV dysfunction, independent of LV involvement. TyG-BMI, a marker of IR, plays a significant role in RV dysfunction, with the inflammatory factor (hs-CRP) acting as a partial mediator. Among the components of TyG-BMI, FPG emerged as the most significant contributor to RV dysfunction. MR further confirmed a causal relationship between FPG and RV function, independent of TG, BMI, and hypertension.

## Acknowledgments

We are grateful to all the patients who participated in this study. Special thanks to the teams behind the IEU OpenGWAS and GWAS Catalog databases, as well as the developers of the R software packages (*glmnet*, *TwoSampleMR*, *MVMR*, and others), for providing essential tools and resources that facilitated our data analysis.

## Funding

We are a self-funded research project with no budget.

## Disclosures

None.

## Data availability

The datasets and analysis codes used in this study are available from the corresponding author or the first author upon reasonable request.

## Supplemental Material

Supplement Table 1-2.

Supplement Figure 1-2

Supplementary Results

## Notes

### Competing Interest Statement

The authors have declared no competing interest.

### Clinical Trial

This study is a cross-sectional study and thus was not registered on a clinical trial platform; all participants provided informed consent, and the study received rigorous ethical approval from the Ethics Committee of the First Affiliated Hospital of Dalian Medical University.

